# INDEPENDENT ASSOCIATION OF METEOROLOGICAL CHARACTERISTICS WITH INITIAL SPREAD OF COVID-19 IN INDIA

**DOI:** 10.1101/2020.07.20.20157784

**Authors:** Hemant Kulkarni, Harshwardhan V. Khandait, Uday W. Narlawar, Pragati G. Rathod, Manju Mamtani

## Abstract

Whether weather plays a part in the transmissibility of the novel COronaVIrus Disease-19 (COVID-19) is still not established. We tested the hypothesis that meteorological factors (air temperature, relative humidity, air pressure, wind speed and rainfall) are independently associated with transmissibility of COVID-19 quantified using the basic reproduction rate (R_0_). We used publicly available datasets on daily COVID-19 case counts (total n = 108,308), three-hourly meteorological data and community mobility data over a three-month period. Estimated R_0_ varied between 1.15-1.28. Mean daily air temperature (inversely) and wind speed (positively) were significantly associated with time dependent R_0_, but the contribution of countrywide lockdown to variability in R_0_ was over three times stronger as compared to that of temperature and wind speed combined. Thus, abating temperatures and easing lockdown may concur with increased transmissibility of COVID-19.

## INDTRODUCTION

As the novel COronaVIrus Disease-19 (COVID-19) continues to devastate the world, there remains a myriad of unknowns about its pathogenesis, population dynamics, epidemiology, prevention and treatment. Since its introduction into the global susceptible population SARS-CoV-2, the causative agent of COVID-19, has presented several conundrums. It was initially believed that like many other viruses, SARS-CoV-2 may also be responsive to the environmental influences posed by climatic and meteorological factors^1-4^. However, current understanding of the potential role of weather on the spread of SARS-CoV-2 is far from clear.

The COVID-19 outbreaks have been generally more severe in the countries located in the mid-latitudes where the temperature is considerably low in contrast to the tropical countries. Several studies around the world have attempted to specifically establish a relationship between COVID-19 transmission and various meteorological factors.^5-7^ For example, a study conducted in New York, USA, found that mean temperature, minimum temperature and air quality had a significant association with the COVID-19 pandemic.^5^ Similarly, Shi et al^8^ reported a statistically significant correlation between daily temperature and daily count of COVID-19 cases in China and suggested that temperatures above 8–10 °C would lead to a decline in the number of infected cases. In a parallel investigation, Prata et al^5^ concluded that a rise in 1 °C temperature would result in a decrease in the number of daily confirmed COVID-19 cases in Brazil. There have been very few investigations from India in this regard^9-11^ – a country with second largest population size after China. These studies from India have generally indicated a potential role of weather conditions in the spread of COVID-19.

On the other end of the spectrum, a study conducted by Yao et al^12^ concluded that there is no association of COVID-19 transmission with temperature or UV radiation in Chinese cities. Indeed, an elegant, evidence-based review by Brassley et al^6^ summarized the existing evidence in this regard and observed that a. cold and dry conditions may facilitate the spread of the novel coronavirus (2019-nCoV) b. much of the emerging data for 2019-nCoV has yet to be peer-reviewed and is thus needed; and c. relying on weather changes alone to slow the transmission of COVID-19 are unlikely to be sufficient. Considering these recommendations; the variability in the observed associations; and a relative lack of such studies from India, we conducted this investigation on a nationwide sample of geographical locations across India. The primary goal was to test the putative association of geo-meteorological characteristics with rates of COVID-19 transmission and to test its independence from other socio-behavioral interventions like lockdowns and mobility.

## MATERIALS AND METHODS

### Data sources

We selected a total of 46 geographical locations across India. For each selected location (either a city, union territory or district), we collected data for a three-month period (March 1, 2020 through May 31, 2020). Following data items were collected for each study location: daily number of confirmed COVID-19 cases, meteorological data, demographic data and overall geographic data. The meteorological data included 3-hourly recordings of temperature, relative humidity, air pressure, wind speed and rainfall. The demographic data included the 2011 census population and the geographical data included area and elevation. The area and population records were combined to estimate the community density. Lastly, temporal data on the lockdown implementation phases and the mobility of the population (estimated anonymously from the cellphone use data) was collected to study the potential temporal concurrence with COVID-19 transmission.

All data used in this study are publicly available and are completely anonymized. The study was approved by the Institutional Ethics Committee of Government Medical College, Nagpur, India. Following were the sources of data: number of daily COVID-19 cases – https://api.covid19india.org/; meteorological data –https://www.tutiempo.net/ and https://www.worldweatheronline.com/; 2011 census data – https://censusindia.gov.in/2011-common/censusdata2011.html; and geographical data – combination of census data and search on Wikipedia® (https://en.wikipedia.org/wiki/Wikipedia). Lastly, the temporal mobility data was downloaded from the publicly available repository: https://www.google.com/covid19/mobility/. These indicated percent change from baseline mobility on visits to the following five destinations - retail and recreation, grocery and pharmacy, parks, transit stations and workplaces.

### Quantification of COVID-19 transmissibility

Using the daily case count data we estimated the basic reproduction rate (R_0_) in two different ways. First, we estimated the average R_0_ over the entire duration of 92 days period of data collection. For this, we used two methods – the exponential growth (EG) and the maximum likelihood (ML). Second, we estimated the daily R_0_ in a time-dependent (TD) fashion. All estimates of R_0_ require a knowledge of serial interval, the time difference between onset of symptoms in an infector and an infectee. We assumed a gamma distributed serial interval with a mean of 3.96 days and a standard deviation of 4.75 days as reported by Du et al.^13^ We used the R package R_0_ ^14^ to derive all the estimates of R_0_. Finally, we considered the possibility of biased estimates of R_0_ owing to the relative lack of testing facilities, especially during the initial period of the epidemic. For this, we used the method of Lachmann et al^15^ that considers South Korea as the reference country and estimates the degree of undertesting by combining demographic and vital statistics data. Using this method, we derived the possible undertesting on each study day.

### Statistical analysis

Our analyses used estimates of R_0_ as the dependent variable and the geo-meteorological and socio-behavioral characteristics as the explanatory variables. To compare groupwise means we used the Mann-Whitney U test or Kruskal-Wallis test as appropriate. Significance of heterogeneity across study locations was statistically tested using the Q test. Time series data were smoothed using a five-day sliding window technique. Further, to make the different time series (each meteorological characteristic) comparable, we converted them to a series of z-scores. To test the temporal concurrence, we used the cross-correlation between two time series (Pearson’s correlation). To test the association of time series variables with estimated time dependent R_0_, we used multivariable, ordinary least squares regression. Starting with the full model, we conducted stepwise, backward elimination regression modeling with a probability retention criterion of 0.05. Lastly, to quantify the relative contribution of each covariate with time dependent R_0_, we estimated the proportional reduction in error (PRE) using the approach of Judd, McCleland and Ryan.^16^ PRE was estimated as reduction in the residual sum of squares by including a covariate in the full model. Statistical analyses were conducted using the Stata 14.2 statistical package (Stata Corp, College Station, TX). Type 1 error rate of 0.05 was used for hypothesis testing.

## RESULTS

### Representativeness of the study locations

We included 46 locations across India that contained 32 cities, 12 districts and 2 union territories. Figure 1 shows the geographical spread of these locations and the geographic and demographic details for these locations are provided in Supplementary Table 1. The study locations varied widely in terms of the area (range 4.86 – 6039 sq. miles), elevation (range 3 – 11500 feet above sea level) and population density (range 33.6 to 56812.3/sq. mile). The selected locations are distributed across India and represent majority of the states / union territories of India. Meteorological data was available on all the selected study locations.

**Figure 1.**
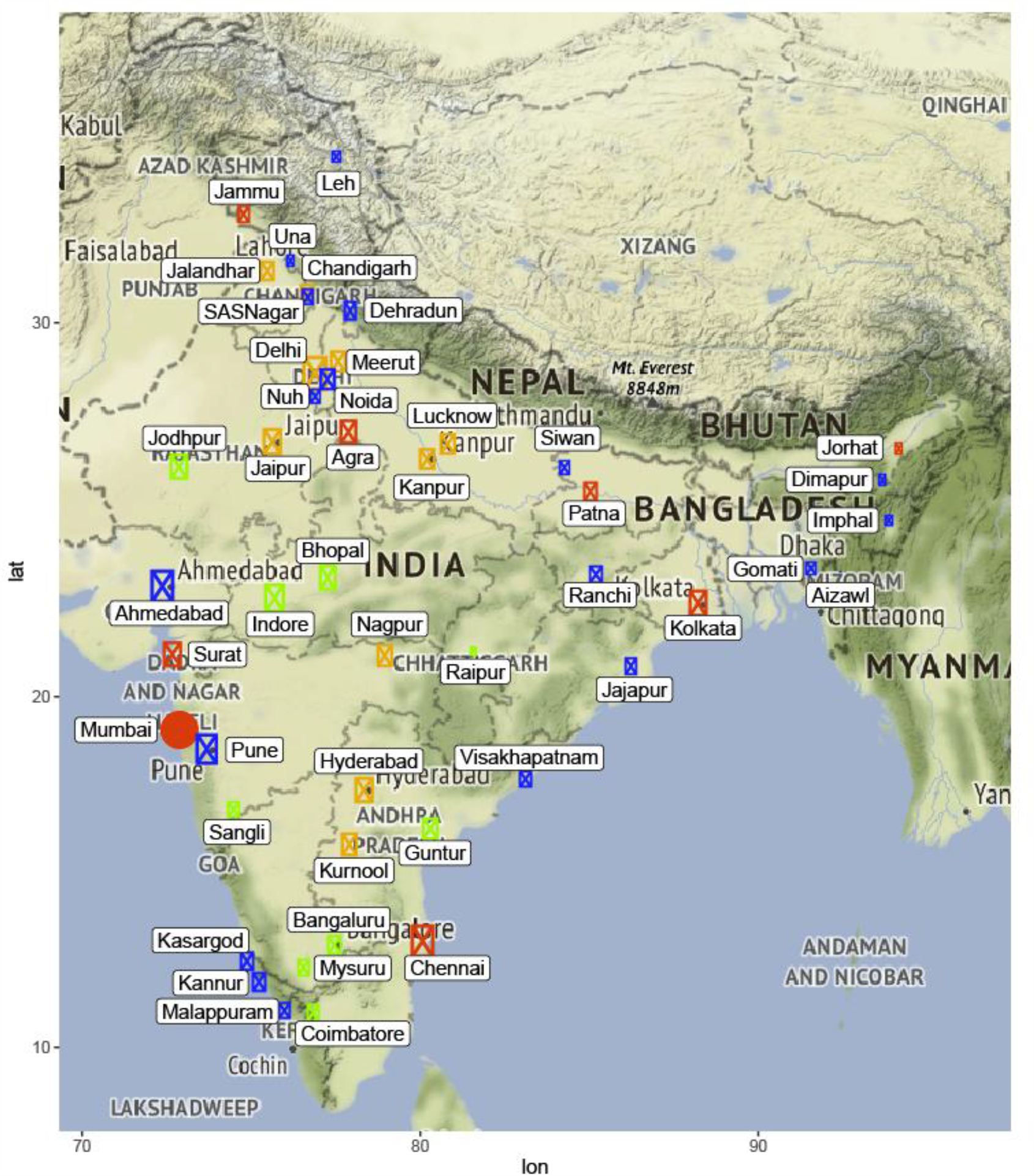
Geographical spread, COVID-19 case counts and population density of the study locations. Selected locations are shown as bubbles, the size of which is proportional to log of COVID-19 case counts. The color of the bubble indicates quartile of population density based on the cutoffs mentioned in Supplementary Table 1 – first quartile, blue; second quartile, green, third quartile orange and fourth quartile, red.

The cumulative number of COVID-19 confirmed cases (till and including May 31, 2020) reported from these locations also varied widely (1 to 37666). The 46 selected locations together accounted for a total of 108,308 confirmed COVID-19 cases. From entire India the number of cumulative COVID-19 cases till May 31, 2020 were 182,140. Thus, our selected geographic locations accounted for ∼60% of all India COVID-19 cases till May 31, 2020. The top 5 contributing locations to the overall cumulative COVID-19 case counts were Mumbai (37666), Delhi (18058), Chennai (12040), Ahmedabad (11919) and Pune (7459) as shown in Supplementary Figure 1.

### Average estimated R_0_ for COVID-19

We first estimated the R_0_ based on case counts reported for the entire country as well as only for the locations included in this study. For each of these datasets, we estimated the R_0_ in two ways – first based on the actual reported case counts and second by inflating the case counts to account for the potential undertesting on each day. The results of these analyses are shown in Figure 2 and referred to as unadjusted (actual case counts, blue bars) and adjusted (for potential undertesting, purple bars). Our average estimates of R_0_ using different methods of estimation and with or without adjusting for undertesting ranged from 1.18 to 1.27 for India and 1.15 to 1.28 for the selected study locations. All the estimates and their 95% confidence intervals (error bars in Figure 2) were significantly above unity. These results indicated that over the study period, the average estimates of R_0_ were significantly greater than one, confirming the existence of the epidemic; the average R_0_ estimates were only moderately above unity; the average R_0_ estimates were minimally influenced by potential undertesting; and that the study locations yielded average R_0_ estimates consistent with those for the whole country thereby indirectly reaffirming the representativeness of the selected study locations.

**Figure 2.**
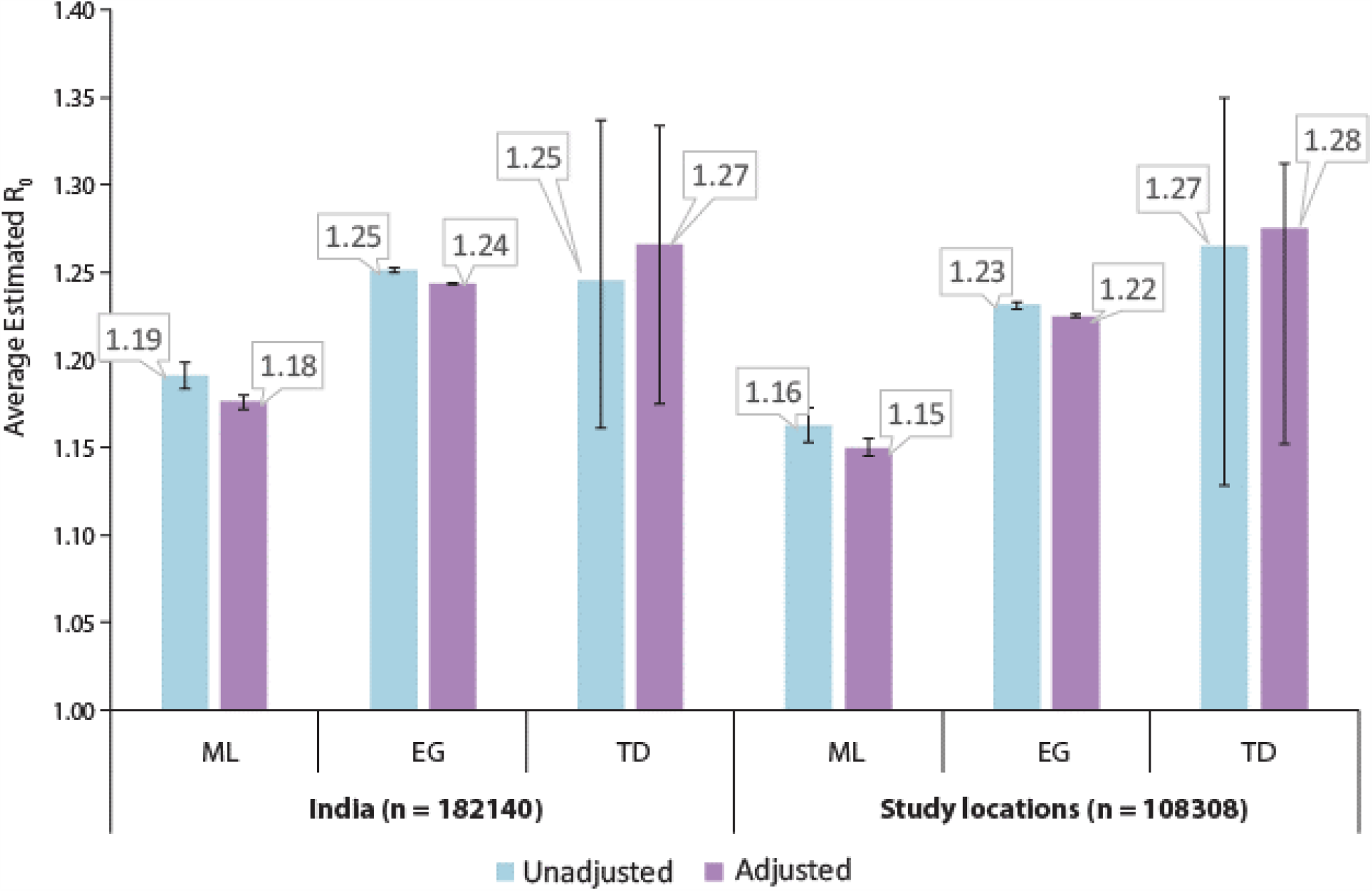
Average estimated R_0_ for COVID-19. Bars show the average R_0_ estimates and error bars indicate the 95% confidence intervals. Average R_0_ estimates were derived using three methods: ML, maximum likelihood; EG, exponential growth; and TD, time dependent. Each estimate was also derived without adjustment (unadjusted, blue bars) and adjusted for potential undertesting (adjusted, purple bars).

We also examined the heterogeneity of the average R_0_ estimates across the study locations. For these analyses, we restricted the locations which showed at least seven consecutive days with a contiguous segment of non-zero cases. Total of 35 locations were eligible based on this criterion. The average R_0_ estimates derived using the ML method [point estimates and confidence intervals (CI)] for these 35 locations are shown in Figure 3. There was a significant heterogeneity in the average R_0_ estimates (p = 6.9×10^−30^) with estimates ranging from 1.98 for Dehradun to 0.89 for Kolkata. The average R_0_ estimates for the top five contributing locations were: Mumbai 1.16 (95% CI 1.14 – 1.18); Delhi 1.25 (95% CI 1.23 – 1.28); Chennai 1.20 (95% CI 1.17 – 1.23); Ahmedabad 1.10 (95% CI 1.07 – 1.13) and Pune 1.22 (95% CI 1.18 – 1.26).

**Figure 3.**
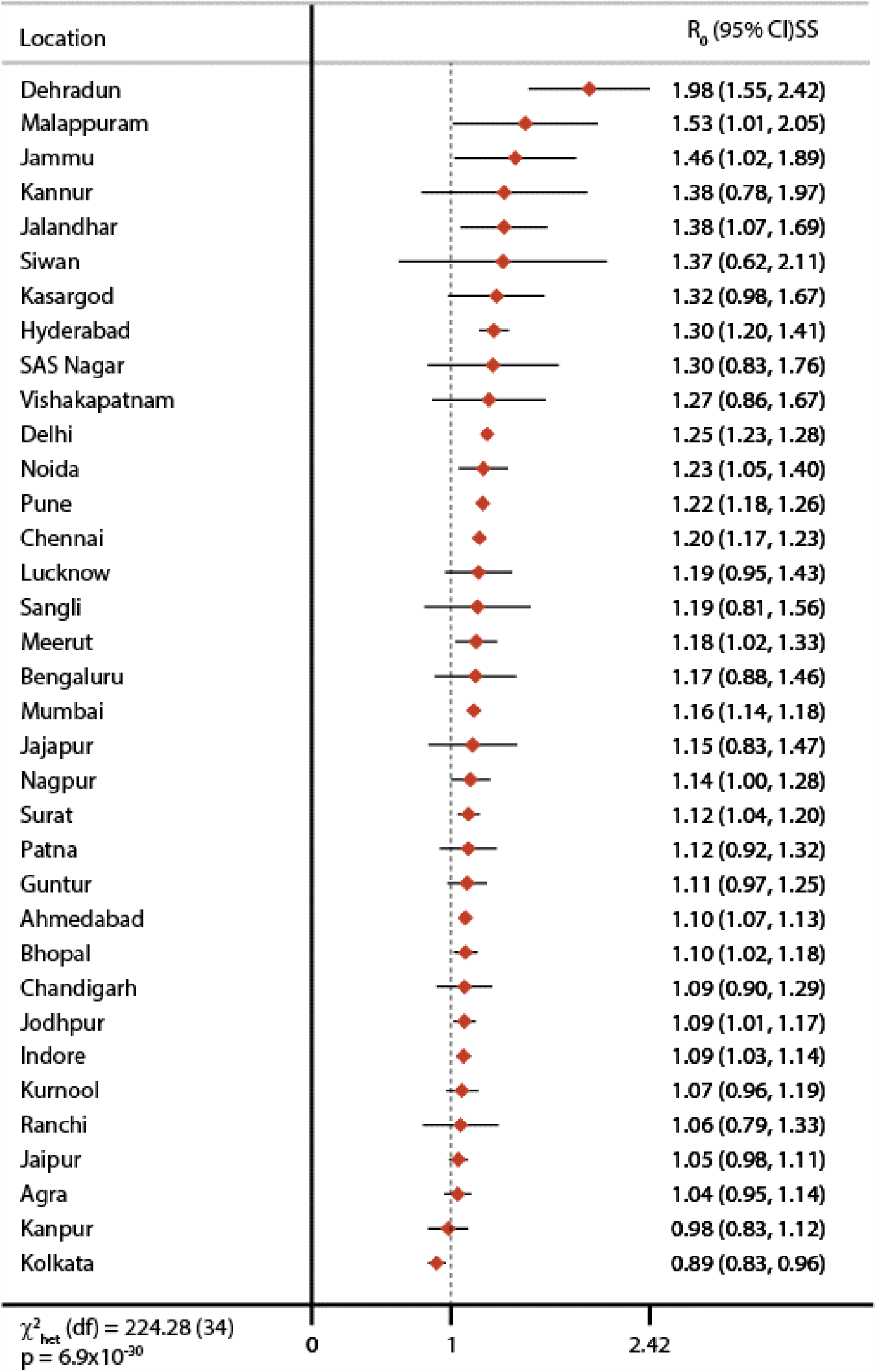
Heterogeneity of R0 estimates across study locations. The forest plot shows point (diamonds) and 95% confidence interval (error bars) estimates for maximum likelihood estimates of R_0_.

### Temporal changes in R_0_ estimates

Next, we considered the variability in R_0_ estimates over the duration of the study for all locations together. Figure 4 shows that the R_0_ estimates were initially high but undulated widely and gradually converged towards the overall estimates shown in Figure 2 with narrow confidence bands later. Thus, the time dependent R_0_ estimates showed considerable variation across study time.

**Figure 4.**
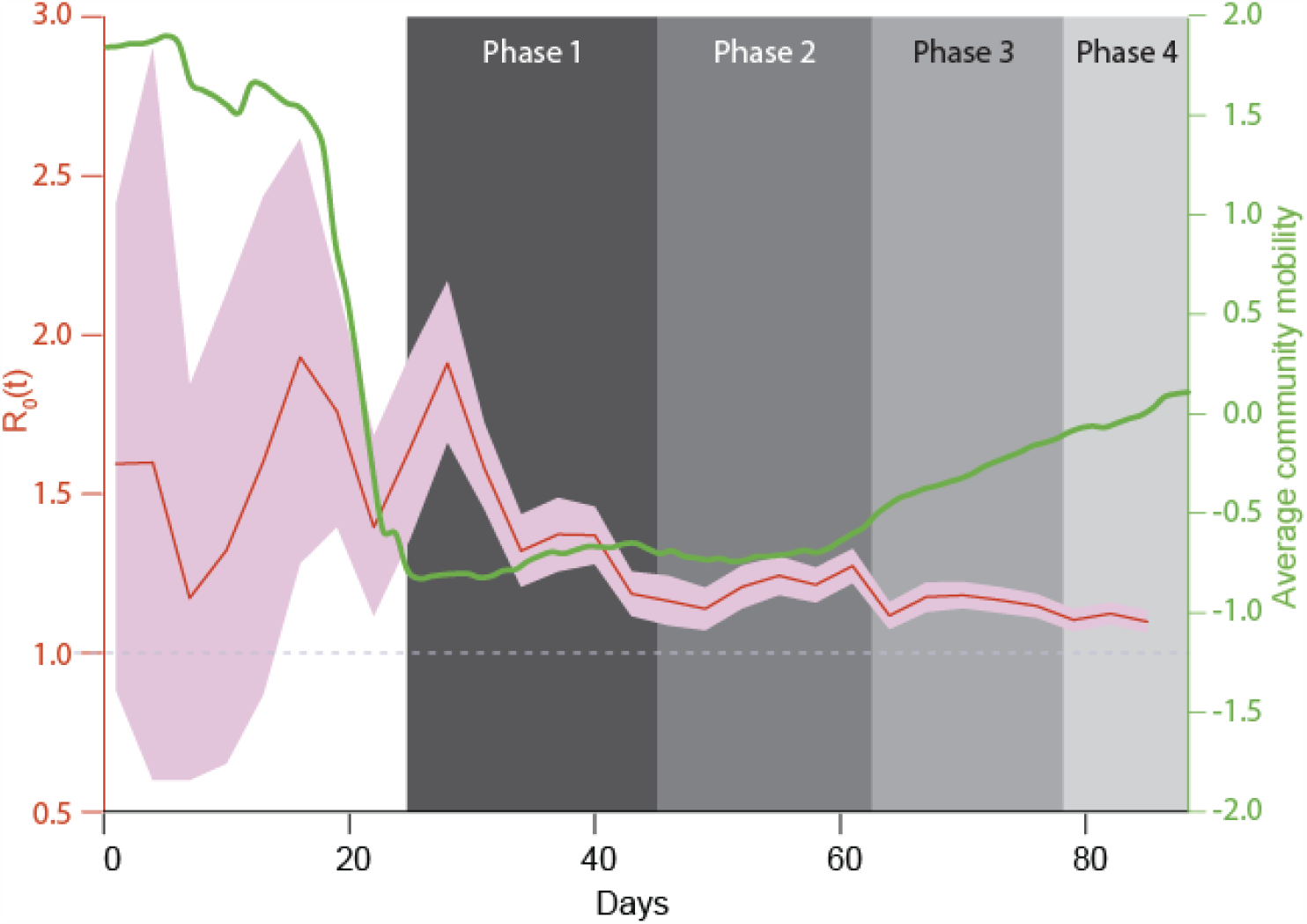
Time dependent R_0_ and socio-behavioral interventions. Red line and pink bands indicate the time dependent R_0_ and 95% confidence intervals, respectively, for each day during the study. These align to the left axis (colored red). The green curve shows the 5-day rolling average z-score for cellphone-based mobility data and aligns to the right axis (colored green). Shaded boxes in the background indicate different phases of the countrywide lockdown in India.

We examined the association of the time-dependent R_0_ estimates with two socio-behavioral characteristics – implementation of a countrywide lockdown and the extent of social distancing as reflected by the cellphone mobility data. When contrasted against the various phases of countrywide lockdown in India (grey shaded regions in Figure 4), we found that the median R_0_ estimates consistently reduced as lockdown was imposed. Before lockdown began (March 1 through March 24, 2020) the median R_0_ estimate was 1.54 and this estimate decreased to 1.40 (March 25 – April 14, 2020), 1.21 (April 15 – May 3, 2020), 1.16 (May 4 – May 17, 2020) and 1.10 (May 18, 2020 onwards) during lockdown phases 1 through 4, respectively (Kruskal-Wallis p <0.0001).

The cellphone-based community mobility data also revealed consistent and interesting patterns. As shown in Supplementary Figure 2, the overall trends in community mobility for all five destinations showed a dramatic decrease around the beginning of phase 1 lockdown, remained very low during phase 1 lockdown and then gradually increased as the lockdown progressed. The 5-day rolling z-scores for the average mobility based on these five parameters is shown in Figure 4 (green curve).

### Association of time dependent estimates with meteorological data

The time trends for temperature, relative humidity, air pressure, wind speed and rainfall are shown in Figure 5A.Over the duration of the study, temperature and wind speed steadily increased; relative humidity and air pressure gradually decreased while rainfall remained steady. As a first step of the association analyses, we estimated the cross-correlation between each meteorological variable and the R_0_ estimates. Figure 5B shows the cross-correlograms for lags ranging from -10 to 10 days. We found that higher temperature, wind speed and rainfall were correlated inversely while relative humidity and air pressure were correlated positively with time dependent R_0_ estimates. The best cross-correlation was observed for temperature and humidity on the same day (-0.73 and 0.63, respectively), wind speed on previous day (-0.40), rainfall preceding by 4 days (-0.29) and air pressure preceding by 6 days (0.54). Together these results indicated that concurrent or immediately preceding values of meteorological variables are significantly correlated with time dependent R_0_ estimates.

**Figure 5.**
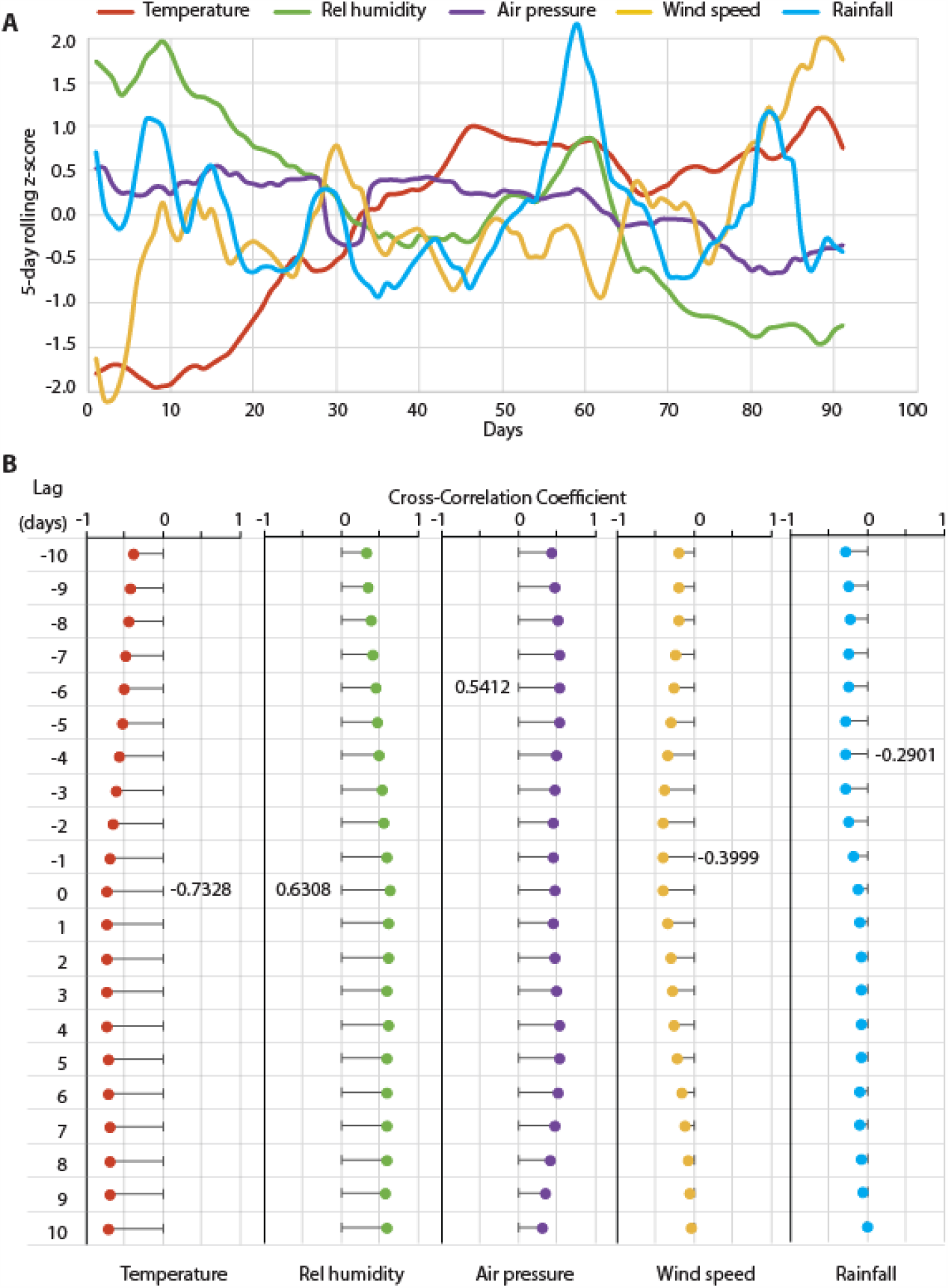
Meteorological determinants of COVID-19 transmissibility. (A) Time trends for each of the five, color-coded meteorological variables. For each variable, the data were first z-transformed and then subjected to a 5-day moving average. (B) Cross-correlograms for correlation of each meteorological variable with estimated time dependent R_0_. All cross-correlograms were assessed between lags of -10 to 10 days. Most significant correlation for each variable is indicated as a number alongside the lag at which it was observed. Rel, relative.

### Multivariable association of meteorological and socio-behavioral predictors with time dependent R_0_

We then examined whether the meteorological and socio-behavioral covariates were independently associated with time dependent R_0_ estimates. The full regression model used time dependent R_0_ estimates as the dependent variable and following 14 covariates as explanatory variables: five z-scores for the meteorological covariates, five z-scores for community mobility data and four phases of lockdown (each used as a dichotomous variable). The results of these analyses are shown in Table 1. In the full model, we observed that the lockdown phases 3 (only marginally) and 4 and wind speed were the only covariates that were statistically significantly associated with R_0_ estimates. In this context, the mobility data (which was highly correlated with the lockdown phases) did not retain statistical significance. However, considering the potential for interactions among covariates and the possibility of an underpowered full model (14 covariates observed on 92 days), we conducted stepwise regression modeling with a probability retention criterion of 0.05. The results of the final model (Table 1) showed that temperature z-scores, wind speed and lockdown phases 2-4 were retained in the final model. This model fitted the data well with an adjusted R^2^ of 0.56 (Supplementary Figure 3)

**Table 1.** Multivariable association of meteorological and socio-behavioral covariates with time dependent R0 estimates (all study locations, March 1 – May 31, 2020)

| Covariate | $\beta$ | 95% CI | p |
| --- | --- | --- | --- |
| FULL MODEL |  |  |  |
| Temperature z-score | -0.10 | -0.24 - 0.05 | 0.190 |
| Relative humidity z-score | -0.04 | -0.16 - 0.09 | 0.539 |
| Air pressure z-score | -0.06 | -0.18 - 0.06 | 0.322 |
| Wind speed z-score | 0.06 | 0.00 - 0.11 | 0.036 |
| Rainfall z-score | 0.02 | -0.03 - 0.07 | 0.471 |
| Retail/recreation z-score | 0.54 | -0.30 - 1.37 | 0.204 |
| Grocery/pharmacy z-score | 0.17 | -0.08 - 0.41 | 0.184 |
| Parks z-score | -0.26 | -0.91 - 0.39 | 0.426 |
| Transit station z-score | -0.48 | -1.18 - 0.23 | 0.180 |
| Workplaces z-score | 0.04 | -0.19 - 0.28 | 0.724 |
| Lockdown phase 1 | -0.01 | -0.27 - 0.25 | 0.936 |
| Lockdown phase 2 | -0.23 | -0.61 - 0.16 | 0.247 |
| Lockdown phase 3 | -0.41 | -0.87 - 0.05 | 0.079 |
| Lockdown phase 4 | -0.61 | -1.14 - -0.08 | 0.024 |
| Intercept | 1.56 | 1.30 - 1.81 | <0.001 |
| FINAL MODEL |  |  |  |
| Temperature z-score | -0.08 | -0.13 - -0.03 | 0.005 |
| Wind speed z-score | 0.08 | 0.03 - 0.12 | 0.003 |
| Lockdown phase 2 | -0.22 | -0.34 - -0.09 | 0.001 |
| Lockdown phase 3 | -0.32 | -0.45 - -0.19 | <0.001 |
| Lockdown phase 4 | -0.47 | -0.63 - -0.30 | <0.001 |
| Intercept | 1.51 | 1.44 - 1.58 | <0.001 |

$\beta$ , regression coefficient; CI, confidence interval; p, significance value

From the point of public health relevance, we then quantified the contribution of each variable retained in the final model to the overall variance of time dependent R_0_. The PRE estimates for the variables retained in the final model were as follows: temperature: 9.1%, wind speed: 9.9%, lockdown phase 2: 12.2%, lockdown phase 3: 22.5% and lockdown phase 4: 27.0%. These results indicate that while the meteorological factors of temperature and wind speed were statistically significant predictors of COVID-19 transmissibility, their contribution to dampening the R_0_ estimate was 3-4 times weaker as compared to the countrywide lockdown phases 2-4.

## DISCUSSION

Using nationwide data from India over a three-month period, our study made three cardinal observations. First, the average basic reproduction rate (R_0_) of COVID-19 infection in the period from March 1 through May 31, 2020 ranged from 1.15 to 1.28 even after accounting for the potential undertesting. Second, the COVID-19 transmissibility (quantified using R_0_) was significantly associated with daily average temperature (inversely), daily average wind speed (positively) and the countrywide intervention of lockdown (inversely). Third, the contribution of lockdown to the variability in time dependent R_0_ was three times more than the contribution of temperature and wind speed combined. Together, these results suggest that in India while the meteorological determinants of COVID-19 were independently associated with the transmissibility, their contribution was outweighed by that of the countrywide lockdown.

Even though statistically significantly greater than unity, our estimate of R_0_ was low. This estimate is comparable to the value of 1.32 reported by Du et al. However, the low value of R_0_ should be interpreted with caution. First, there has been a debate about the length of serial interval with values ranging from as low as 3 days to as high as 9 days.^13, 17-21^ We used the serial interval of ∼4 days which is on the lower side of the serial interval range and could have partly contributed to the low R_0_ observed in this study. Second, the major part of the study period included lockdown and reduced mobility and therefore the R_0_ estimate may represent a muted transmissibility owing to interventions in place. Third, the low R_0_ estimate does not indicate lack of viral infectiousness or any other viral characteristic but only implies the extent of potential spread of the disease.^22^ Fourth, the epidemic of COVID-19 is still ongoing and our estimate of R_0_ only captures the initial, ascending limb of the epidemic curve. Therefore, this R_0_ estimate does not fully capture the population dynamics of COVID-19. Fifth, our estimate of R_0_ is a conglomerate of the varying estimates across the study locations as shown in Figure 3. The variability in R_0_ across study locations indicates that the location-specific epidemic curves were not aligned to the same starting point in time and therefore our R_0_ estimate should not be used as a generalizable estimate of COVID-19 transmissibility. The reason for estimating R_0_ in the study was to investigate the potential influence of geo-meteorological factors on transmissibility.

Several researchers around the world have demonstrated an inverse relationship between air temperature and COVID-19 transmissibility.^23-27^ Our results also are in agreement with the general understanding that higher ambient temperature can inversely influence COVID-19 transmissibility.^23, 27^ The study duration mark a period of increasing temperature in the Indian peninsula and our results indicate that, in general, high ambient temperatures were associated with lower R_0_ estimates such that unit standard deviation increase in air temperature was associated with a 0.08 lower R_0_ (Table 1, final model).On the other hand, we observed that a unit standard deviation increase in wind speed was associated with a 0.08 higher R_0_ (Table 1, final model). The current evidence for the potential role of wind speed in COVID-19 spread is conflicting with studies reporting positive,^28^ null^29-31^ and negative^1, 32^ association with COVID-19 transmissibility. Our observation of a positive association of COVID-19 transmissibility with wind speed is in line with the growing idea that the SARS-CoV-2 virus may be airborne.^33, 34^ Of note, incidence of COVID-19 has been shown to be associated with air pollution^1, 35, 36^ – a factor that is significantly influenced by wind speed.^37^ Our study cannot directly answer these interesting hypotheses, which should be tested in future studies. Nevertheless, a head-to-head comparison indicated that the lockdown period was associated with three times stronger contribution to the variability in R_0_ as compared to that of air temperature and wind speed combined. From the perspective of public health action, this observation supports the role of proactive interventions to de-escalate the transmissibility of COVID-19. Conceivably, as the temperatures wane and the lockdown is eased, more cases of COVID-19 can be expected.

Our results should be interpreted in the light of some limitations. First, this was a retrospective analysis that combined data from different sources. The data are collected at the level of geographic locations and not at the level of individual patient. For example, person-to-person transmissibility of COVID-19 in an infector-infectee scenario was not investigated in this study. Therefore, all the estimates and associations should only be considered as general patterns rather than definitive evidence. Second, akin to any observational study, unmeasured confounding can be expected to be operational. Despite these potential limitations our study demonstrated interesting and important patterns of association of geo-meteorological factors in COVID-19 spread. To control a pandemic of this magnitude, all scientific evidence from a holistic standpoint is needed. To that end, our study provides clues into the ecological aspects of COVID-19 during the initial months in India.

## Data Availability

All data used in the study are publicly available.

https://api.covid19india.org/

https://www.tutiempo.net/

https://www.worldweatheronline.com/

https://censusindia.gov.in/2011-common/censusdata2011.html

https://en.wikipedia.org/wiki/Wikipedia

https://www.google.com/covid19/mobility/

## Author Bio

Dr. Kulkarni is the Chief Executive Officer of M&H research, LLC, in San Antonio, Texas, USA and is also the President of the not-for-profit Lata Medical Research Foundation based in Nagpur, India. Dr. Kulkarni has published over 130 papers on various aspects of biomedical research including non-communicable diseases, HIV and AIDS, genetics, environmental sciences and mathematical modeling of infectious diseases.

